# Awareness, knowledge, attitude and practice towards measures for prevention of the spread of COVID-19 in the Ugandans: A nationwide online cross-sectional Survey

**DOI:** 10.1101/2020.05.05.20092247

**Authors:** Robinson Ssebuufu, Franck Katembo Sikakulya, Simon Mambo Binezero, Lucien Wasingya, Sifa K. Nganza, Bwaga Ibrahim, Patrick Kyamanywa

## Abstract

**Background:** The world is facing the Coronavirus pandemic which is highly infectious. A number of measures have been put in place to prevent its spread among the population. However, for these preventive measures to be effective, the population requires an appropriate and sufficient knowledge. Thus, a survey was conducted with the aim of assessing the awareness, knowledge, attitude and practices towards measures for prevention of the spread of Covid-19 amongst Ugandans.

**Methods:** This was a cross-sectional study conducted during the lockdown in Uganda. An online questionnaire and a snowballing approach was used for participant recruitment of 18 years above literate Ugandans. Data collection was done from 6^th^ to 15^th^ April 2020 during which 1763 people participated. We analyzed all data using STATA 14.2, applying appropriate statistical tests.

**Results:** Out of 1763 participants, 97.6% were aware of the current pandemic. 83.9% of participants had a good knowledge score (21.8/27), 72.4% had a good attitude and 85.3% were practicing measures to prevent the spread of the Coronavirus disease. The ordered logistic regression showed that being a Health worker was significantly associated with a high knowledge (aOR:6 (3.32-10.93); a good attitude (aOR:2.5(1.68-3.8)]) and good practice (aOR:2.9 (1.95-4.2). On contrary, being a driver, business entrepreneur and a security personnel were found to have less rate in awareness, knowledge, attitude and practice.

**Conclusions:** Ugandans had a good overall awareness, knowledge, attitude and practice. However, there is still a gap of knowledge, attitude and practice among drivers, business entrepreneur and security personnel. There is a need to mobilize the population in the country to have the same degree of awareness and knowledge which will have an impact on attitude and practice to prevent spread of COVID-19.

## 1. Introduction

On 31st December 2019, a respiratory syndrome identified to be caused by a beta-coronavirus was reported in Wuhan, China [1]. This was later officially named as an outbreak of a new coronavirus disease-2019 (COVID-19) by World Health Organization(WHO) [2], and as the severe acute respiratory syndrome coronavirus 2 (SARS-CoV-2) by Coronavirus Study Group (CSG) of the International Committee on Taxonomy of Viruses, on 11 February 2020 [3],[4].

Due to the rapid global spread of this disease, the COVID-19 was declared as a pandemic on 11th March 2020 [5]. SARS-CoV-2 presents clinically with fever, dry cough, fatigue, myalgia, and dyspnea [5]; [6]. The COVID-19 virus is transmitted between people through droplets, fomites and close contact, with possible spread through the eyes, nose and mouth but it is not an airborne disease according to the current studies [7]. The disease is highly contagious with enormous potential for health, economic and societal impacts [6].

As of 28^th^ April 2020, a total of 2, 954,222 cases of COVID-19 had been confirmed worldwide (22,376 confirmed in Africa) with 202, 597deaths (899 deaths registered in Africa) giving a crude fatality ratio of 6.9% worldwide (4 % in Africa) [7], [8].

Several community-based and facility-based measures have been put in place to contain the spread of the Coronavirus and its impact on the health systems and populations. The key community-based measures include self-isolation, hand-washing with soap, restriction of movements with lock down measures, sanitization of surfaces. Facility-based measures have so far included use of personal protective equipment before handling patients, testing of symptomatic patients, treatment and contact tracing, in addition to quarantine of the suspected and diagnosed cases [8];[9]. The success of such measures however is dependent on the adherence by the population, a factor of their awareness, knowledge, attitude and behavior.

Uganda registered her first case of COVID-19 on 21^st^ March, 2020 and as of 28^th^ April 2020, according to the Ministry of Health (MOH) information, Uganda had registered 79 confirmed cases of covid-19 with 52 recoveries and without any reported death to date (https://www.health.go.ug). COVID-19 is a rapidly evolving disease and currently, there is neither vaccine nor evidence on the effectiveness of potential therapeutic agents [3].

Given the spread of the new coronavirus and its impacts on human health, the WHO has recommended strategies to control this pandemic disease which include traffic restriction, cancellation of social gatherings, home quarantine, establishment of clinical care and management strategies, laboratory capacity strengthening, surveillance strategies, case and contact tracing, infection prevention and control, implementation of health measures for travelers, risk communications and community engagement [8];[9].

These measures can be achieved through awareness within the population and if the population has appropriate knowledge and correct attitude on the transmission and how to prevent spread of COVID-19. To our knowledge, there is no study yet done in Africa and Uganda in particular, to evaluate the level of awareness, the knowledge and attitudes of the population about the transmission and preventive measures put in place to mitigate the COVID-19 outbreak. However, previous studies on viral disease outbreaks, like SARS in 2003[10] and Ebola in 2018(12), have shown that the management and control of an outbreak requires a good understanding of the populations about the disease to avoid its spread in the community [6]. It is therefore necessary that a survey is undertaken to establish the level of awareness, the knowledge and attitudes of the population with regard to the COVID-19 pandemic and the measures put in place to mitigate it.

The continent of Africa has poorly-equipped health settings to manage thousands of COVID-19 infected people in comparison to developed countries. It is also clear from the current reports that even the health-care systems in the high-income countries have been overwhelmed by patients in spite of the fact that they are better equipped. Therefore, the best strategy for a low-resourced setting like Africa and Uganda in particular would be to mitigate the spread by quickly improving the awareness, knowledge and attitude and the adherence of the population to the preventive measures in place. However, there is a paucity of data on public knowledge and attitudes of prevention of COVID-19. It was, therefore important that a survey like this be carried out so that evidence-based strategies are put in place to address shortcomings identified.

This survey aimed at assessing the awareness, knowledge and attitudes towards measures for prevention of the spread of Covid-19 among the Ugandans.

## 2. Materials and Methods

### 2.1 Study design and setting

This was a nationwide cross-sectional online survey conducted on the Ugandans using the snowball sampling technique.

### 2.2 Study participants

All literate Ugandans aged 18 years and above constituted the population of this survey. The population of Uganda stands at 44,269,594 which is 78.4% of the literate Ugandans [12]. In Uganda, around 20 million people have a mobile subscription, representing 44% of the general Ugandans among whom nearly half of all mobile subscribers also access mobile internet services [13]. By June 2018, there were nearly 10 million mobile internet connections in Uganda a penetration rate of 23% [13].

All literate Ugandans with a minimal computer literacy level assumed to be able to operate at least an email, WhatsApp, Tweeter or Facebook and also consented to participate were included in the survey. Those who had filled in the form but for some reason were unable to submit the questionnaire were automatically not reflected and therefore excluded in the data base for the survey.

### 2.3 Data collection and instrument

For a period of ten days from 6^th^ to 15^th^ April 2020, an online snowball sampling technique was used with a semi-structured questionnaire which was developed using Google forms (https://docs.google.com/forms/d718Ded-NFe65B6HnTFCwe4XzxBA-o3VfcJYogRjgcMsAA/edit), with a consent form appended to it. The link of the questionnaire was sent through emails, WhatsApp and other social media to the contacts of research assistants. The participants were encouraged to roll out the survey to as many people as possible. Thus, the link was forwarded to other people apart from the first point of contact. On receiving and clicking to the link the participants were auto directed to the information about informed consent and the survey. After reading the preamble and consent part of the questionnaire, they were directed to participate in the survey that started with demographic details followed by a set of several questions on knowledge, attitude and practice that appeared in that sequence.

The questionnaire was composed of 19 items and developed based on WHO requirements for KAP (knowledge, attitudes and practices) [14];[6] and focused on several key constructs: (1) socio-demographics characteristics (age, sex, profession, location, and marital status); (2) Awareness about Covid-19 and affective responses of the participants about Covid-19;(3) Knowledge about the measures putted in place to prevent the spread of Covid-19; (4) attitudes towards the mitigating measures putted in place; (5) practice towards the mitigating measures putted in place.

Questions related to Awareness about Covid-19 and affective responses of the participants about Covid-19 involved “whether covid-19 exists; covid-19 is curable, covid-19 leads to death and participants are worried about covid-19.

Knowledge about the measures put in place to mitigate the spread Covid-19 which involved the incubation period of Covid-19; the signs or symptoms of Covid-19; Probable origin of transmission of covid-19; the risks factors for severe illness of Covid-19; measures to control the spread covid-19, Prevention, hand washing and face mask.

A correct answer was assigned 1 point and an incorrect/unknown answer will be assigned 0 points. The total score ranged from 0 to 27, with a higher score denoting a better/high attribute of COVID-19. 27 points scoring as: 0-9 as poorly knowledgeable, 10-18 as moderately knowledgeable and 19-27 as highly knowledgeable.

Attitudes towards the measures put in place to mitigate spread of covid-19 included *“staying home and call for help from the hotline that has been provided by Ministry of Health or District of health if having signs or symptoms of covid-19”* was interpreted as Agree (Yes) and Disagree ( No/Going to hospital).

Practice towards the measures put in place to mitigate spread of covid-19 which involved “*measures observed for Self-Prevention”* was scored as bad practice for 0-4points; fair practice for 4-6 points and good practice for the total of 7 points.

### 2.4 Data processing and analysis plan

The raw data was cleaned and entered into Microsoft excel and exported into STATA 14.2 for processing. Statistical analysis was done using STATA 14.2, where by categorical variables were summarized using frequency tables and graphs while continuous variables were summarized using means and standard deviation (SD). We did bi-variate analysis between the knowledge score, attitude, practice and Awareness/Affective groups and social demographic characteristics using the Chi-square. For bi-variate analysis between the knowledge score and social demographic characteristics using the T-TTEST and ANOVA for variables with two and more than two groups respectively. Finally, multiple analysis was also done using ordered logistic and logistic regressions for ordered score groups and bivariate variables respectively reporting adjusted odds ratios.

### 2.5 Ethical considerations

Ethical clearance for the survey was obtained from the Institutional Research Ethical Committee of Kampala International University in Uganda (UG-REC-023/201914). As participant logged in online, a statement regarding the consent to participate in the survey was on the top of the questionnaire. and it was assumed that all participants could only proceed after reading the consent and accepting to participate in the survey.

The participation in this survey was voluntary. Participants were free to withdraw from the survey at any time by not submitting their form online and there was no any repercussion. The participants’ identity was concealed as the form does not require any identification. No name or mail was required from the participant. Therefore, the information obtained was a onymous and was stored anonymously and treated confidentially. Only the 5 members research team was allowed to access data and only the principal investigator accessed the entire dataset. There was no human risk associated with this survey, as we were not meeting participants in person and no names, contact information or biological samples were obtained from the participants.

It our hope that the findings of this survey will be of benefit to Ugandans in exploring and understanding their awareness, knowledge and attitudes on prevention of COVID-19 and this enhanced risk communication to inform efforts on how to mitigate the spread of the disease.

## Results

A total of one thousand seven hundred sixty-eight (1768) participants completed the online questionnaire. Five (5) participants were excluded from the survey because were aged below 18years, thus the final sample size considered was one thousand seven hundred sixty-three (1763).

**Table 1:**
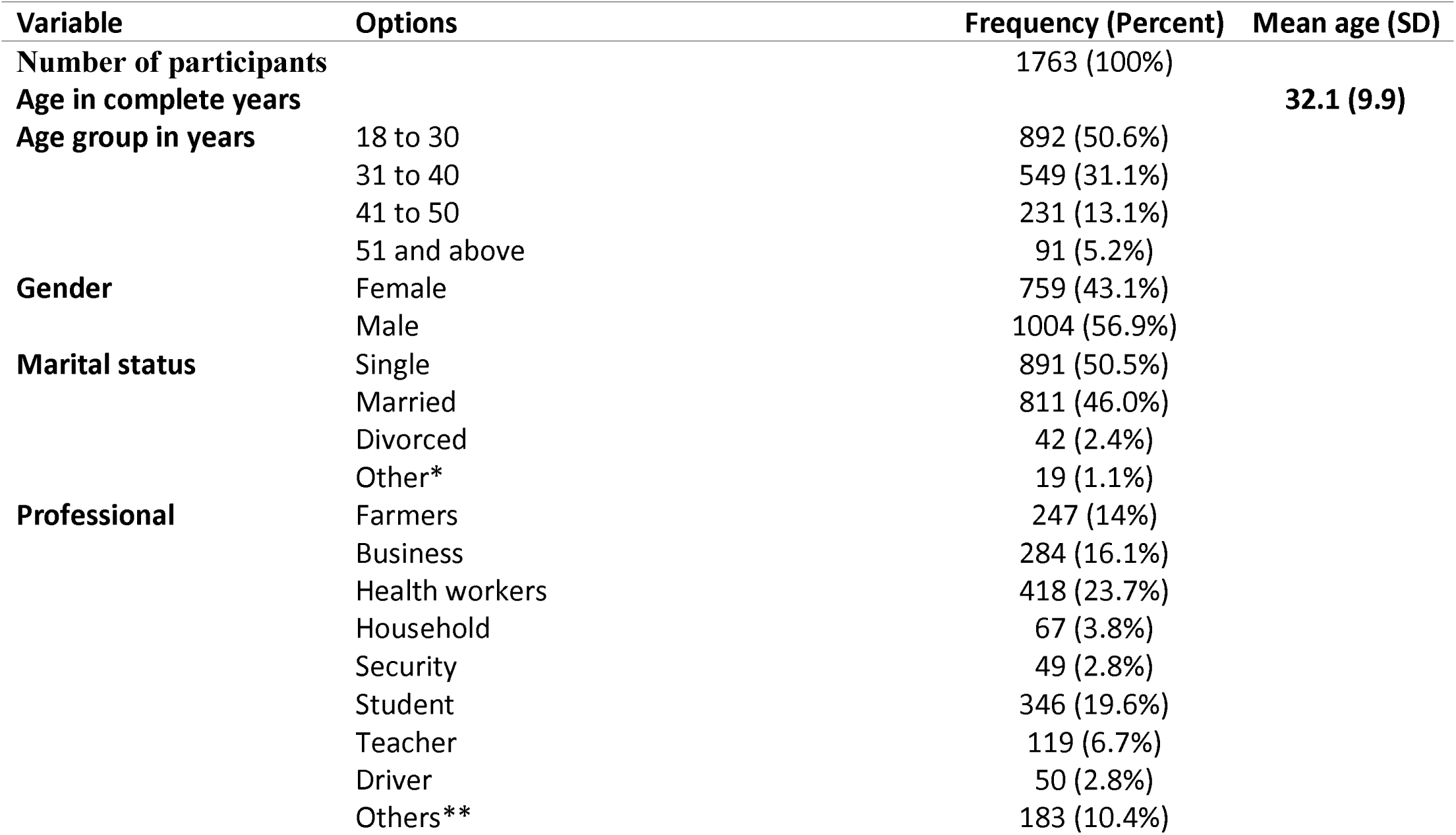
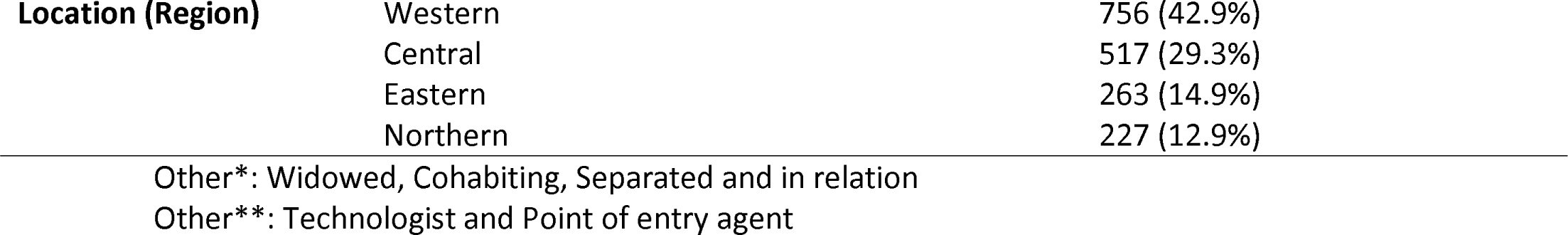
Socio-demographic characteristics of participants. Out of 1763 participants, 56.9% were male and 50.5% were single. The mean age of the overall respondents was of 32.1(± 9.9) years. 23.7% were health workers, 14% farmers and 2.8% drivers. The majority (42.9%) of participants were from Western region of Uganda, followed by Central Uganda (29.3%). Other socio-demographic characteristics are shown in Table 1.

| Variable | Options | Frequency (Percent) | Mean age (SD) |
| --- | --- | --- | --- |
| **Number of participants** |  | 1763 (100%) |  |
| **Age in complete years** |  |  | **32.1 (9.9)** |
| **Age group in years** | 18 to 30 | 892 (50.6%) |  |
|  | 31 to 40 | 549 (31.1%) |  |
|  | 41 to 50 | 231 (13.1%) |  |
|  | 51 and above | 91 (5.2%) |  |
| **Gender** | Female | 759 (43.1%) |  |
|  | Male | 1004 (56.9%) |  |
| **Marital status** | Single | 891 (50.5%) |  |
|  | Married | 811 (46.0%) |  |
|  | Divorced | 42 (2.4%) |  |
|  | Other* | 19 (1.1%) |  |
| **Professional** | Farmers | 247 (14%) |  |
|  | Business | 284 (16.1%) |  |
|  | Health workers | 418 (23.7%) |  |
|  | Household | 67 (3.8%) |  |
|  | Security | 49 (2.8%) |  |
|  | Student | 346 (19.6%) |  |
|  | Teacher | 119 (6.7%) |  |
|  | Driver | 50 (2.8%) |  |
|  | Others** | 183 (10.4%) |  |

|  |  |  |
| --- | --- | --- |
| **Location (Region)** | Western | 756 (42.9%) |
|  | Central | 517 (29.3%) |
|  | Eastern | 263 (14.9%) |
|  | Northern | 227 (12.9%) |

Other\*: Widowed, Cohabiting, Separated and in relation
Other\*\*: Technologist and Point of entry agent

The Figure 1 shows that most Ugandans (97.6%) had heard about the ongoing coronavirus pandemic. Although, 73.3% of participants considered the covid-19 to be curable, 26.7% of them thought otherwise. 97.0% of participants thought that covid-19 can lead to death and 10.9% of participants were worried about the ongoing pandemic. (**Fig 1**)

**Figure 1:**
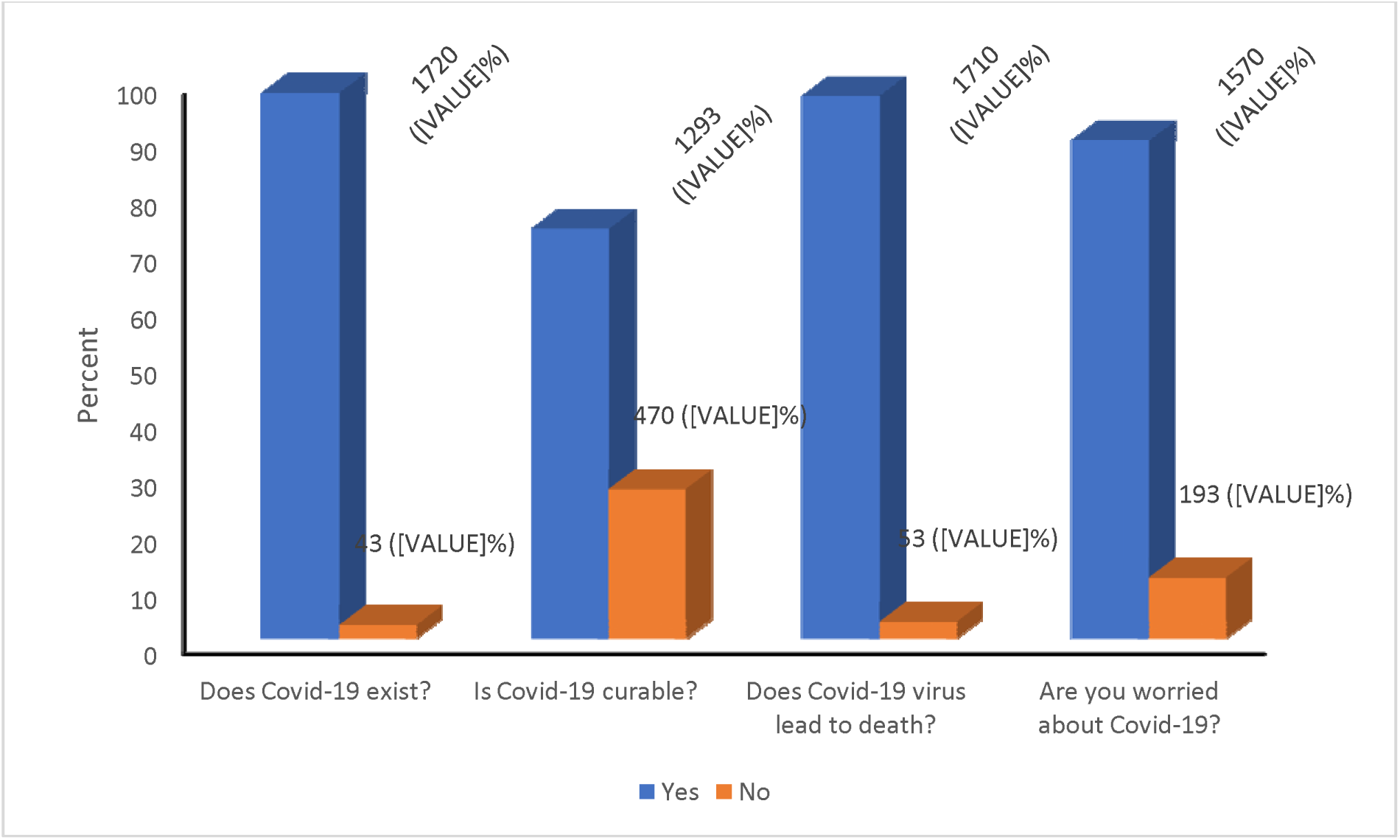
Awareness / Affective Responses of participants.

**Table 2:** Knowledge score means (SD) with socio-demographic characteristics of participants. Table 2 shows Knowledge Score Means (SD) with Socio-demographics characteristics of participants with a mean of COVID-19 knowledge score of 21.8 (±4.0) suggesting overall 80.7% (21.8/27*100) correct rate on this knowledge test.

| Variable | Sample Size | Knowledge score (SD) | P-Value |
| --- | --- | --- | --- |
| **Total** | 1763 | 21.8 (4.0) |  |
| **Age group in years** |  |  |  |
| 18 to 30 | 892 | 22.0 (3.9) | 0.113 |
| 31 to 40 | 549 | 21.8 (3.9) |  |
| 41 to 50 | 231 | 21.5 (4.7) |  |
| 51 and above | 91 | 21.2 (4.1) |  |
| **Gender** |  |  |  |
| Female | 759 | 21.3 (4.5) | 0.001 |
| Male | 1004 | 22.2 (3.6) |  |
| **Marital status** |  |  |  |
| Single | 891 | 22.1 (3.8) | 0.001 |
| Married | 811 | 21.7 (4.1) |  |
| Divorced | 42 | 19.0 (5.6) |  |
| Others* | 19 | 21.7 (4.8) |  |
| **Professional** |  |  |  |
| Farmers | 247 | 18.5 (5.1) | 0.001 |
| Business | 284 | 20.8 (4.0) |  |
| Health workers | 418 | 23.8 (2.4) |  |
| Household | 67 | 19.8 (5.1) |  |
| Security | 49 | 21.0 (3.9) |  |
| Student | 346 | 22.6 (3.1) |  |
| Teacher | 119 | 22.7 (2.9) |  |
| Driver | 50 | 18.2 (4.8) |  |
| Other** | 183 | 23.4 (2.6) |  |
| **Location (Region)** |  |  |  |
| Western | 756 | 21.7 (4.0) | 0.003 |
| Central | 517 | 21.6 (4.7) |  |
| Eastern | 263 | 22.5 (3.0) |  |
| Northern | 227 | 22.3 (3.4) |  |

Other\*: Widowed, Cohabiting, Separated and in relation
Other\*\*: Technologist and Point of entry agent

## Discussion

As per now the world is facing the coronavirus pandemic which is highly infectious, measures have been put in place to prevent its spread among the population across the world. The population requires an appropriate and sufficient knowledge about these measures, their importance and how to apply them appropriately [8];[9]. To our knowledge, there is no survey yet done in Sub-Saharan Africa and Uganda in particular to evaluate the level of understanding of African population about the transmission and preventive measures put in place to mitigate the COVID-19 outbreak. Therefore, it was imperative to conduct a survey among African population where disaster could be expected as the health setting in this continent are not well equipped to manage thousands of people infected with coronavirus disease.

When a human population is facing an outbreak, changes in behavior in response to the disease can alter the progression of the infectious agent. In particular, people aware of a disease in their proximity can take measures to reduce their susceptibility [15].

However, beyond a critical infection rate, spreading awareness can slow down the spread of the disease and lower the final incidence, but it cannot completely stop it from reaching epidemic proportions and taking over large parts of the population [6]. Only if the disease is easily recognized and information spreads rapidly, while at the same time there is a strong tendency toward protective behavior, awareness of a disease outbreak can bring the infection rate of a disease down significantly. If all of these factors work together, rapid drops in the transmissibility of a disease, as have been observed in the 2003 outbreak of SARS in Hong Kong[16].

Our survey reported that most of Ugandans were aware of the ongoing coronavirus pandemic (97.6%). Despite that awareness, 73.3% of participants considered that the covid-19 is curable, though 26.7% of them rejected that notion (**Figure 1**). 10.9% of participants were worried about the ongoing pandemic with a high thought that covid-19 to lead to death (97.0%).

These findings are different from a survey done in USA. The survey looked at covid-19 among adults with chronic conditions, in which all participants reported having heard about the coronavirus, although 24.6% of the participants of that survey were worried about the current pandemic [17]. Action is needed to increase awareness of Ugandans to reduce the observed gap in awareness and to ensure that all population are adequately made aware of the gravity of the COVID-19 pandemic.

In India, it was found that despite the high level of awareness among adult population about coronavirus pandemic, there was an increase in worries and apprehensions among the public regarding acquiring of the COVID-19 infection which indicated a need of intensifying the awareness program among Indian population and address the mental health issues of people during this COVID-19 pandemic [18].

As of 28^th^ April 2020, a cumulative total of 22, 376 confirmed COVID-19 cases with 899 deaths (case fatality ratio: 4%) have been reported across the 45 affected countries in Africa continent including Uganda [8].

Ten days after the first case of Covid-19 being confirmed in Uganda, we conducted an nationwide online survey on awareness, knowledge, attitudes and practice towards measures for prevention of the spread of covid-19 among Ugandan population, we found that the overall participant had 80.7% on knowledge towards measures for prevention of the spread of Covid-19 in the Ugandans. This result is approximately similar to the knowledge rate (90%) found among Chinese residents during a quick online survey on covid-19 [6] and during the Ebola outbreaks in Sierra Leone in 2014 [19] and DRC in 2018 [11]. This result could be explained by the fact that the covid-19 found Ugandans already familiar with observing similar measures to prevent the spread of some other highly infectious diseases within the country such as Ebola and Marburg disease. The ordered logistic regression from our survey showed us that the level of knowledge was significantly associated with a certain degree of education level as per Health worker [aOR:6 (3.32-10.93)], teacher [0R:5.9 (2.64-13.29)] and Student [aOR:3.3 (1.9-5.64), p<0.001]. As this survey included all Ugandan with a minimal computer literacy level constituted 78.4% of all the Ugandan population [12] and in particular those who are able to operate an email, WhatsApp, Tweeter, Facebook where the Ministry of Health is currently posting information related to measures for prevention of the spread of the pandemic within the country, this could explain the findings mentioned above and which could be different among uneducated people. Zhong *et al*. findings related to knowledge in China were explaining their findings by the fact that the most repondents during their survey held an associate’s degree or higher [6]. The Uganda government could use these categories of participants as a strategy to reach out and sensitize the uneducated population about measures to be observed among all Uganda.

Uganda is surrounded by countries having many cases of covid-19 and this can expose its population to be infected by the ongoing pandemic. This case been shown by truck drivers from Tanzania and Kenya who have tested positive to COVID-19 (https://www.health.go.ug). Most respondents (72.4%) agreed that you stay at home and call for help from the hotline that has been provided by Ministry of Health or District health Officer if they have symptoms or signs of covid-19. This finding can be explained by the high percent of knowledge among participants and also the country is under lockdown as one of measures to prevent the spread of the pandemic. In Chine, a survey revealed that most population took precautions to prevent infection by COVID-19 such as not going to crowded places and wearing masks when going outside but with an optimistic attitudes towards COVID-19 which could be attributed to the very strict prevention and control measures implemented by local governments such as banning public gatherings In Nigeria, a study on Ebola outbreak evaluating the KAP of Ebola virus infection among secondary school children, found an association between inadequate knowledge and carried a negative attitude towards the outbreak [20]. In our survey, householder [aOR: 1.2 (0.67-2.28)] and driver profession [aOR: 1.5 (0.73-2.89)] were associated with a poor attitude. During the Ebola outbreak in DRC and Guinea, it was found during these studies that attitude of a group of participants had poor attitude towards measures for prevention Ebola in their areas [11];[21] the same as for the study in Chinese residents during this coronavirus pandemic [6]. Among confirmed cases in Uganda, one third of them seem to be drivers who are coming from surrounding countries. This result gives us the useful information that government has to increase sensitization among these categories of people about measures towards prevention of the spread of covid-19 which can be considered as a cross border infection in the Uganda and East African region. Household can get information from the students and those who have shown a high knowledge about measures to observed and the government can pass through such a category of people to have her strategies to control the pandemic within the country to be observed among all Ugandans. Our result Despite having a high knowledge (80.7%), good attitude (72.4%) and good practice in 85.3%( together good and fair practice) among participants which is much better than what found during a study on knowledge and attitude towards Ebola and Marburg virus outbreaks in two affected communities in Uganda [22], 14.7% of participants were not practicing social distancing; hand washing; self-monitoring; use of Masks; cleaning surfaces; avoid gatherings; stay at home. Being a driver [aOR: 1.2 (0.66-2.1)], householder [aOR: 1.5 (0.88-2.67)], security agent [aOR: 1.3 (0.7-2.43)], doing business [aOR: 1.2 (0.85-1.74)] and those located were found to be associated with poor practice measures of prevention of the spread of covid-19. This result can be explained by the fact that participants within these professions had poor knowledge in ordered logistic regression analysis (Table 3) compare to the one expected having a certain level of education and being in Northern Uganda was associated with poor attitude (Table 4).

**Table 3.** Ordered logistic regression of Knowledge score means and group with socio-demographic characteristics of participants. The mean Knowledge scores significantly differed across genders, marital status, profession and location (P<0.05) but did not significantly differ across age groups (p>0.05) in univariate analysis and ordered logistic regression analysis as shown in Table 3; The different sources of information about covid-19 among participants were as follows; social media in 36.8% followed by television (29.9%), health-workers (12.9%), radio (12%), Family and friends (5.2%) and News Paper (3.2%). Out of 1763 participants, 1479 (83.9%) scored 19-27 and were therefore highly knowledgeable about covid-19 followed by 257 (14.6%) moderately knowledgeable and 27 (1.5%) poorly knowledgeable. The ordered logistic regression of knowledge score (Table 3) shows that being a Health worker [aOR:6 (3.32-10.93), P<0.001]; teacher [0R:5.9 (2.64-13.29, p<0.001]; Student [aOR:3.3 (1.9-5.64), p<0.001] was significantly associated with a high knowledge score.. On contrary, being a business merchant [OR: 1.5 (0.96-2.27), p:0.078]; security agent [aOR: 1.9 (0.83-4.28), p:0.131] and driver [0.5 (0.26-1.05),p: 0.069] was associated with poor knowledge about covid-19. Participants residing in the East and Northern regions of Uganda were associated with a high knowledge. [aOR: 3.4 (2.06-5.77)] and [aOR: 1.9 (1.14-3.26)] respectively. However, being in Central Uganda was associated with poor knowledge [aOR: 1.3 (0.87-1.87)].

| Variable | Knowledge Score Mean (SD) |  | Knowledge Score group |  |  |
| --- | --- | --- | --- | --- | --- |
|  | Coefficient (95%CI) | P-Value | Coefficient (95%CI) | aOR (95%CI) | P-Value |
| **Age group in years** |  | **0.371** |  |  | **0.566** |
| 18 to 30 | Ref |  | Ref |  | Ref |
| 31 to 40 | 0.3 (-0.09-0.75) | 0.121 | 0.2 (-0.17-0.63) | 1.3 (0.84-1.88) | 0.260 |
| 41 to 50 | 0.4 (-0.15-0.97) | 0.150 | 0 (-0.51-0.5) | 1 (0.6-1.65) | 0.982 |
| 51 and above | 0.4 (-0.39-1.16) | 0.329 | -0.1 (-0.73-0.59) | 0.9 (0.48-1.81) | 0.832 |
| **Gender** |  | **0.027** |  |  | **0.171** |
| Female | Ref |  | Ref | Ref |  |
| Male | 0.4 (0.04-0.69) | 0.027 | 0.2 (-0.09-0.53) | 1.2 (0.91-1.7) | 0.171 |
| **Marital status** |  | **0.004** |  |  | **0.048** |
| Single | Ref |  | Ref | Ref |  |
| Married | 0.2 (-0.17-0.64) | 0.249 | 0.2 (-0.2-0.59) | 1.2 (0.82-1.8) | 0.335 |
| Divorced | -1.7 (-2.73--0.59) | 0.002 | -0.9 (-1.79--0.09) | 0.4 (0.17-0.91) | 0.030 |
| Other* | 0.2 (-1.28-1.74) | 0.766 | 0.2 (-1.37-1.84) | 1.3 (0.25-6.3) | 0.775 |
| **Professional** |  | **0.000** |  |  | **0.000** |
| Farmers | Ref |  | Ref | Ref |  |
| Business | 1.3 (0.68-1.85) | 0.000 | 0.4 (-0.04-0.82) | 1.5 (0.96-2.27) | 0.078 |
| Health workers | 3.6 (3.01-4.16) | 0.000 | 1.8 (1.2-2.39) | 6 (3.32-10.93) | 0.000 |
| Household | 1.1 (0.18-1.98) | 0.019 | 0.6 (-0.15-1.25) | 1.7 (0.86-3.5) | 0.122 |
| Security | 1.7 (0.67-2.71) | 0.001 | 0.6 (-0.19-1.45) | 1.9 (0.83-4.28) | 0.131 |
| Student | 2.8 (2.2-3.45) | 0.000 | 1.2 (0.64-1.73) | 3.3 (1.9-5.64) | 0.000 |
| Teacher | 2.9 (2.2-3.68) | 0.000 | 1.8 (0.97-2.59) | 5.9 (2.64-13.29) | 0.000 |
| Driver | -0.4 (-1.39-0.63) | 0.465 | -0.6 (-1.34-0.05) | 0.5 (0.26-1.05) | 0.069 |
| Other** | 3.2 (2.53-3.86) | 0.000 | 1.8 (0.99-2.51) | 5.8 (2.7-12.32) | 0.000 |
| **Location (Region)** |  | **0.000** |  |  | **0.000** |
| Western | Ref |  | Ref | Ref |  |
| Central | 0.6 (0.21-0.99) | 0.003 | 0.2 (-0.14-0.62) | 1.3 (0.87-1.87) | 0.213 |
| Eastern | 1.2 (0.73-1.66) | 0.000 | 1.2 (0.72-1.75) | 3.4 (2.06-5.77) | 0.000 |
| Northern | 0.4 (-0.11-0.87) | 0.131 | 0.7 (0.13-1.18) | 1.9 (1.14-3.26) | 0.015 |

aOR: Adjusted Odds Ratio
Other\*: Widowed, Cohabiting, Separated and in relation
Other\*\*: Technologist and Point of entry agent

**Table 4:** Ordered logistic regression of Attitude towards covid-19 with socio-demographic characteristics of participants. Most of participants (72.4%) agreed versus 27.6% who disagreed that they are staying home and call for help from the hotline that has been provided by Ministry of Health or District of health Officer if they have symptoms or signs of covid-19. However, in the ordered logistic regression Table 4 the attitude significantly differed across profession (p<0.001). Teacher [aOR:2.8 (1.59-4.86)]; health workers [aOR:2.5(1.68-3.8)] and students [aOR: 2.3 (1.51-3.61)] were associated with a good attitude but being a driver [aOR: 1.5 (0.73-2.89)] and householder [aOR: 1.2 (0.67-2.28)] was associated with a poor attitude.

| Variable | Attitude Score group |  | P-Value |
| --- | --- | --- | --- |
|  | Coefficient (95%CI) | aOR (95%CI) |  |
| **Age group in years** |  |  | **0.626** |
| 18 to 30 | Ref | Ref |  |
| 31 to 40 | 0.1 (-0.19-0.43) | 1.1 (0.83-1.53) | 0.441 |
| 41 to 50 | 0.3 (-0.14-0.68) | 1.3 (0.87-1.98) | 0.198 |
| 51 and above | 0.1 (-0.48-0.62) | 1.1 (0.62-1.86) | 0.807 |
| **Gender** |  |  | **0.444** |
| Female | Ref | Ref |  |
| Male | 0.1 (-0.14-0.33) | 1.1 (0.87-1.39) | 0.444 |
| **Marital status** |  |  | **0.196** |
| Single | Ref | Ref |  |
| Married | 0.3 (-0.03-0.56) | 1.3 (0.97-1.74) | 0.081 |
| Divorced | -0.1 (-0.82-0.67) | 0.9 (0.44-1.95) | 0.838 |
| Other* | 0.8 (-0.44-2) | 2.2 (0.65-7.42) | 0.208 |
| **Professional** |  |  | **0.001** |
| Farmers | Ref | Ref |  |
| Business | 0.4 (0.04-0.82) | 1.5 (1.04-2.27) | 0.032 |
| Health workers | 0.9 (0.52-1.34) | 2.5 (1.68-3.8) | 0.000 |
| Household | 0.2 (-0.4-0.82) | 1.2 (0.67-2.28) | 0.501 |
| Security | 0.7 (-0.01-1.45) | 2.1 (0.99-4.27) | 0.053 |
| Student | 0.8 (0.41-1.28) | 2.3 (1.51-3.61) | 0.000 |
| Teacher | 1 (0.46-1.58) | 2.8 (1.59-4.86) | 0.000 |
| Driver | 0.4 (-0.31-1.06) | 1.5 (0.73-2.89) | 0.285 |
| Other** | 0.6 (0.18-1.11) | 1.9 (1.2-3.04) | 0.006 |
| **Location (Region)** |  |  | **0.000** |
| Western | Ref | Ref |  |
| Central | -0.8 (-1.07--0.49) | 0.5 (0.34-0.61) | 0.000 |
| Eastern | -0.6 (-0.93--0.25) | 0.6 (0.39-0.78) | 0.001 |
| Northern | -1 (-1.4--0.7) | 0.4 (0.25-0.5) | 0.000 |

aOR: Adjusted Odds Ratio
Other\*: Widowed, Cohabiting, Separated and in relation
Other\*\*: Technologist and Point of entry agent

Our results were similar to the study done in China; having a good practice was related to being a health worker or student and from their findings the authors suggested that the health education intervention would be more effective in sensitizing the other categories of population about measures to observe prevention of the current coronavirus pandemic [6]. Overall participants about KAP, the profession of security gent, driver and business were having less rate of KAP as indicated above (Tables 3-5). This category of population needs an urgent sensitization across the country to avoid the spread of covid-19. Unfortunately, the government of Uganda has allowed the movement of these people (driver, business entrepreneur and security agents) within the country and across borders during the ongoing lockdown. If measures are not taken under consideration, Uganda could increase its number of confirmed cases by these categories of Ugandans.

**Table 5:** Ordered logistic regression of Practice of measures towards covid-19 with socio-demographic characteristics of participants. 260 (14.7%) participants were not practicing social distancing; hand washing; self-monitoring; use of Masks; cleaning surfaces; avoid gatherings; stay at home, though, the majority of the survey participants 1503 or 85.3% (together good fair practice) was practicing these measures to stop the spread of covid-19. However, the practice of respondents was significantly associated with profession and location of participants in the ordered logistic regression (P<0.001) with being health workers [aOR:2.9 (1.95-4.2)], teacher [aOR: 2 (1.24-3.17)] and students [aOR: 2 (1.34-3)] associated with good practice but being a driver [aOR: householder [aOR: business [aOR: 1.2 (0.85-1.74)] and those located in Northern Uganda [aOR: 0.9 (0.67-1.25)] were associated with poor practice measures of prevention of the spread of covid-19 (Table 5).

| Variable | Practice Score group |  |  |
| --- | --- | --- | --- |
|  | Coefficient (95%CI) | aOR (95%CI) | P-Value |
| **Age group in years** |  |  | **0.057** |
| 18 to 30 | Ref | Ref |  |
| 31 to 40 | 0.3 (0.06-0.62) | 1.4 (1.06-1.86) | 0.018 |
| 41 to 50 | 0.4 (0-0.74) | 1.5 (1-2.1) | 0.050 |
| 51 and above | 0 (-0.47-0.52) | 1 (0.63-1.68) | 0.923 |
| **Gender** |  |  | **0.591** |
| Female | Ref | Ref |  |
| Male | -0.1 (-0.28-0.16) | 0.9 (0.76-1.17) | 0.591 |
| **Marital status** |  |  | **0.137** |
| Single | Ref | Ref |  |
| Married | -0.2 (-0.44-0.1) | 0.8 (0.64-1.11) | 0.221 |
| Divorced | -0.7 (-1.37--0.08) | 0.5 (0.25-0.92) | 0.027 |
| Other* | 0.2 (-0.92-1.33) | 1.2 (0.4-3.77) | 0.723 |
| **Professional** |  |  | **0.000** |
| Farmers | Ref | Ref |  |
| Business | 0.2 (-0.16-0.56) | 1.2 (0.85-1.74) | 0.275 |
| Health workers | 1.1 (0.67-1.43) | 2.9 (1.95-4.2) | 0.000 |
| Household | 0.4 (-0.13-0.98) | 1.5 (0.88-2.67) | 0.136 |
| Security | 0.3 (-0.36-0.89) | 1.3 (0.7-2.43) | 0.408 |
| Student | 0.7 (0.3-1.1) | 2 (1.34-3) | 0.001 |
| Teacher | 0.7 (0.22-1.15) | 2 (1.24-3.17) | 0.004 |
| Driver | 0.2 (-0.42-0.74) | 1.2 (0.66-2.1) | 0.589 |
| Other** | 1.1 (0.66-1.62) | 3.1 (1.93-5.03) | 0.000 |
| **Location (Region)** |  |  | **0.000** |
| Western | Ref | Ref |  |
| Central | 0.5 (0.2-0.74) | 1.6 (1.22-2.1) | 0.001 |
| Eastern | 0.6 (0.26-0.9) | 1.8 (1.3-2.47) | 0.000 |
| Northern | -0.1 (-0.41-0.22) | 0.9 (0.67-1.25) | 0.567 |

Ref: Reference variable group
aOR: Adjusted Odds Ratio
Other\*: Widowed, Cohabiting, Separated and in relation
Other\*\*: Technologist and Point of entry agent

The survey was limited to participants who had smartphones, computers, tablets and with internet connectivity and had an understanding of English. Therefore, those with no smartphones and internet connectivity were unable to access the online form and participate in the survey. The survey captured the literate population of the country, so it could not be generalized to the whole population. The awareness, knowledge and attitudes in uneducated people might be different from the findings of this survey.

Therefore, awareness, knowledge and attitudes towards COVID-19 of vulnerable populations deserve special research attention. In addition to the limited sample representativeness, the other limitation of this survey was the inadequate assessment of attitudes towards COVID-19, which should be developed via focus group discussion and in-depth interview and constructed as multi-dimensional measures. But this was not possible due to the lockdown in the country during the survey period and as one of strategies observed by all population in the country was social distancing to avoid the spread of the COVID-19.

## Conclusion

In summary, we found that most of Ugandans are aware, knowledgeable and have good attitude and observing good practice towards measures to prevent the spread of COVID-19 within the country. Despite these findings, we found that there is a lack of knowledge, attitudes and practice among a certain group of population (driver and security agents). This group should be targeted for sensitization or else they become the source of the spread of the coronavirus disease within the country. There is a dire need of taking action of mobilizing all population around the country to have the same degree of awareness and knowledge which will have an impact on attitude and practice. The government of Uganda could use the health workers, teachers and students to help in mobilization of all population within the country about measures towards prevention of the spread of the coronavirus pandemic in the country.

## Data Availability

The data that were used to obtain the findings are available from the corresponding author FKS and the author RS if needed.

## Acknowledgement

Authors thank Ugandans who participated in the survey and research assistants who cared of this survey during data collection.

## Authors contributions

RS and FKS conceived and designed the survey, supervised the online data collection and critically reviewed the manuscript.

SBM participated in conception of Google data form;

LW and SKN participated in online data collection

PK critically reviewed the manuscript.

---

**S1 Fig. Awareness / Affective Responses of participants**

